# Prediction of the COVID-19 Epidemic Trends Based on SEIR and AI Models

**DOI:** 10.1101/2020.04.21.20074138

**Authors:** Shuo Feng, Zebang Feng, Chen Ling, Chen Chang, Zhongke Feng

## Abstract

The outbreak of novel coronavirus-caused pneumonia (COVID-19) in Wuhan has attracted worldwide attention. To contain its spread, China adopted unprecedented nationwide interventions on January 23. We sought to show how these control measures impacted the containment of the epidemic. We proposed an SEIR(Susceptible-Exposed-Infectious-Removed) model to analyze the epidemic trend in Wuhan and use the AI model to analyze the epidemic trend in non-Wuhan areas. We found that if the closure was lifted, the outbreak in non-Wuhan areas of mainland China would double in size. Our SEIR and AI model was effective in predicting the COVID-19 epidemic peaks and sizes. The implementation of control measures on January 23, 2020, was indispensable in reducing the eventual COVID-19 epidemic size.

## 1. Introduction

Since the end of the 20th century, new respiratory infections [1] have emerged in many parts of the world [2]. Among them, the genus *β*-coronavirus of the coronavirus [3] family poses a continuing threat to human health due to its high transmission efficiency, severe infection consequences, and unpredictable timing of epidemics [4]. Over the past few decades, humans have faced many challenges with viral respiratory infections, including SARS-COV in China in 2002 [5,6], H1N1 [7] in Mexico in 2009 [8] and MERS-COV in Saudi Arabia in 2012 [9,10].

In December 2019, the first case of a 2019 coronavirus patient was found in Wuhan, Hubei Province [11]. On January 23,2020, Chinese government closes off Wuhan [12]. The pathogen was named severe acute respiratory syndrome coronavirus 2 (SARS-cov-2) by the international committee for the classification of viruses on February 11, 2020 [13,14]. The name of the disease caused by SARS-cov-2 is COVID-19. Within two months, COVID-19 had spread [15,16] rapidly from Wuhan to all parts of the country. According to China’s national health commission, by March 6, the total number of confirmed cases was 80,653.

Scholars from various countries have attempted to study and analyze the epidemic situation of COVID-19 by various means [17-19]. On January 24, British scholars Read [20] et al. used the SEIR(Susceptible-Exposed-Infectious-Removed) model to predict the trend of the epidemic. They predicted that the number of infections in Wuhan would reach 190,000 by February 4. This estimate clearly overestimated the trend of the outbreak. On January 27, Biao Tang [21] et al. used epidemic data from January 10 to January 22 to predict the epidemic regeneration coefficient of 6.47 (95% confidence interval 5.71-7.23) by the SEIR(Susceptible-Exposed-Infectious-Removed) model and statistical calculation method. Their model estimated that the number of infections would peak on approximately March 10. In early February, Norden E. Huang [22] et al. proposed a simple data-driven model based on natural growth, predicting that the number of infections would peak on approximately February 5, with a cumulative number of confirmed cases between 37,000 and 44,000. On February 24, 2020, Huwen Wang et al. [23] used SEIR(Susceptible-Exposed-Infectious-Removed) model and proposed that the infection coefficient R0 decreased from 2.5 to 0.5 with virus variation and government policy, which had an impact on the prediction.On 19 March 2020, Joseph T. Wu et al. [24] used SEIR(Susceptible-Exposed-Infectious-Removed) model and they study the influence of infection rate, removal rate in different age groups and migration data on prediction model.On March 25, 2020, Moritz U.G.Kraemer et al.[25] used GLM (generalized linear models) model to consider the impact of population migration and age on the number of infected people.On February 28, Zhong Nanshan et al. [26] used the SEIR model and LSTM model of population migration in Wuhan to predict the trend of the epidemic. The model focused on the impact of population migration in Wuhan on epidemic trends.

In the early stage of the epidemic, due to the lack of sufficient data, it was difficult for scholars to accurately predict the trend of the epidemic [20-23]. Additionally, a number of recent studies have shown that the development of the epidemic is closely related to population movement [24-26]. Therefore, it is still necessary to study the epidemic trend of COVID-19 at the current stage, which is of practical significance for the analysis, prevention and control of the epidemic[27]. According to the characteristics of the Wuhan and non-Wuhan regions, the SEIR model and deep learning model were established, respectively, considering the population flow. By using the actual data and referring to the existing literature and reports, the parameters of the new model were fitted. Finally, the model was used to estimate and analyze the epidemic trend.

## 2. Materials and Methods

### 2.1. Data sources

The epidemic data used in this paper were from the latest epidemiological data of COVID-19 reported by the Ding Xiang Yuan [28]. The urban migration index was derived from the Baidu migration project [29], which is based on the users of Baidu and related products to count and calculate the daily number of pedestrian movements in and out of cities by railway, air and highway. The population density, per capita GDP and other urban data of all provinces in China were obtained from the official website of the National Bureau of Statistics [30]. In this paper, the distance from each province to Wuhan was obtained from the Ovi interactive map [31]. The average temperature of each province in 2019 was from the China Meteorological Administration [32]

### 2.2. SEIR model

In this section, we briefly discuss the properties of the basic Susceptible-Exposed-Infected-Removed (SEIR) model. The model divided people into four categories: Susceptible (S): individuals not yet infected; Exposed (E): individuals experiencing incubation duration; Infectious (I): confirmed cases; Removed (R): recovered and dead individuals.

Figure 1 shows how individuals move through each compartment in the mode.

**Figure 1.**
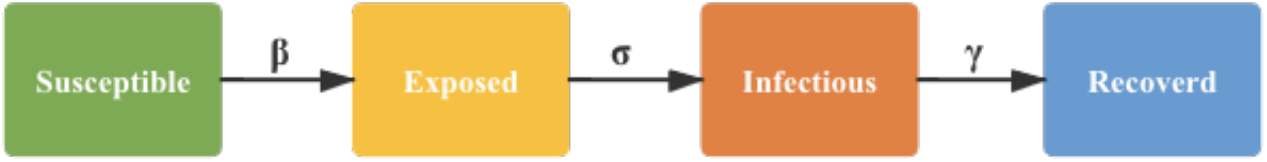
SEIR model with 4 states.

The parameters within this model are as follows:

1. Contact rate *β* controls the rate of spread, which represents the probability of transmitting disease between a susceptible and an infectious individual.
2. Incubation rate *σ* is the rate of latent individuals becoming infectious.
3. Recovery rate *γ* is the rate of infected individuals becoming recovered.

The transmission of the virus is then described by the following system of nonlinear ordinary differential equations:

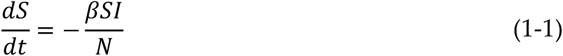

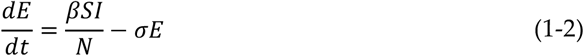

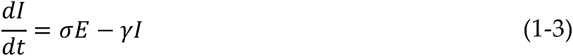

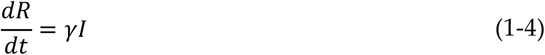

### 2.3. DNN model

In this section, we briefly discuss the properties of the Deep Neural Network (DNN) model. The DNN model first trained a large quantity of data to reduce the loss function and then calculated and updated the parameters in the network to achieve the prediction of new data.

The following equations describe the linear relationship between input and output:

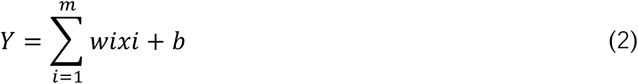

In the equations, *xi* represents the input data, *wi* represents the weight parameter, and b represents the bias.

The DNN model in this study consists of a 9-node input layer, a 32-node hidden layer and a 1-node output layer. Tanh was used as the activation function to increase the fitting degree of the model to the nonlinear model. Additionally, dropout was used in this study to alleviate the overfitting phenomenon of the model. The formula of the Tanh activation function is as follows:

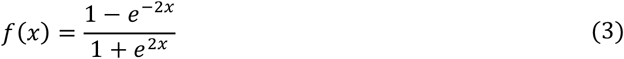

The DNN model architecture used in this study is shown in Figure 2.

**Figure 2.**
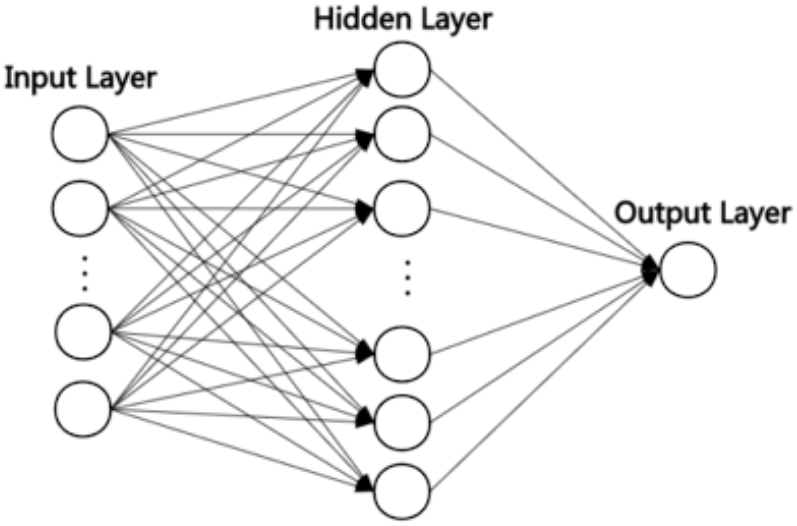
DNN model architecture. The model consists of a 9-node input layer, a 32-node hidden layer and a 1-node output layer.

The loss function can quantitatively determine the quality of the model to select the optimal model. In this study, the mean square error loss function was used. The formula is as follows:

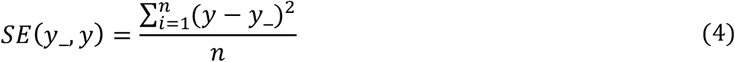

According to the current state for the loss function, we need to update the weight in the direction of the minimum loss to obtain the optimal model. This study uses the Adam (Adaptive Moment Estimation) optimizer to update the weight. Adam is an adaptive learning rate optimization algorithm. By computing the first-moment estimate and the second raw moment estimate of a gradient, an independent adaptive learning rate is designed for different parameters. The calculation formula is as follows:

Update biased first-moment estimate

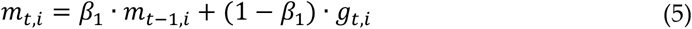

Update biased second raw moment estimate

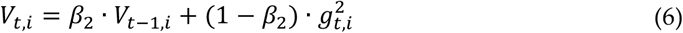

Update parameters

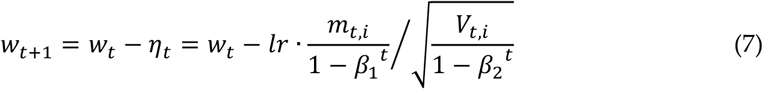

### 2.4. RNN model

In this section, we discuss the use of the Recurrent Neural Network (RNN) is used to predict the number of COVID-19 infections. RNN computes the feature extraction of time-series-based samples by recursion of input sequence data in the direction of sequence evolution.

The RNN consists of input layers, hidden layers, and output layers. The RNN model is shown in Figure 3, and the expansion of Figure 3 is shown in Figure 4.

**Figure 3.**
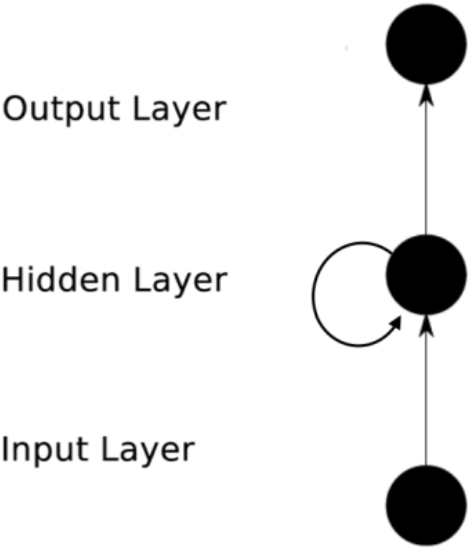
RNN model architecture. The RNN consists of an input layer, a hidden layer, and an output layer.

**Figure 4.**
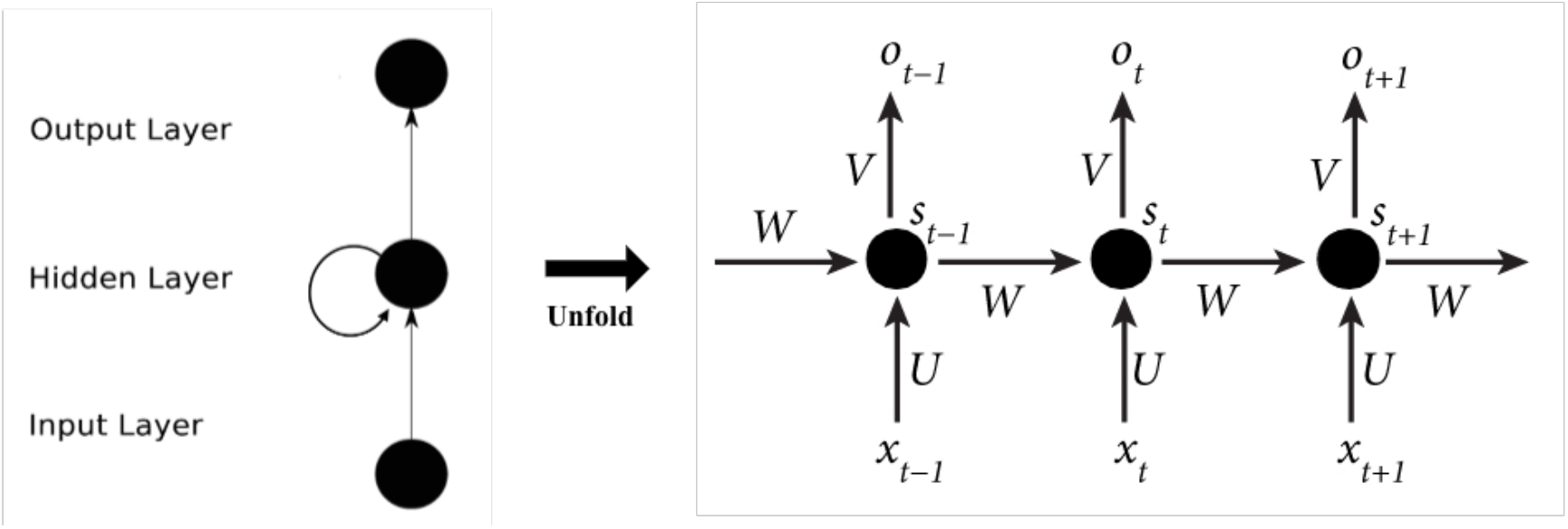
Expansion of the RNN. *t* − 1, *t, and t* + 1 are time series. The input units are {*x*_0_, *x*_1_, …, *x*_*t*_, *x*_*t*+1_, …}, and the output units are {*o*_0_, *o*_1_, …, *o*_*t*_, *o*_*t*+1_, …}. The hidden units are {*s*_0_, *s*_1_, …, *s*_*t*_, *s*_*t*+1_, …} There is a unidirectional flow of information from the input units to the hidden units and another unidirectional flow of information from the hidden units to the output units. W is the weight of the input, U is the weight of the input units at the moment, and V is the weight of the output units.

The calculation formula is as follows:

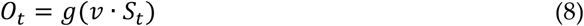

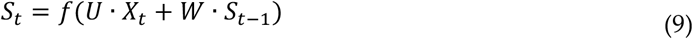

f and g are activation functions.

In this study, two layers of the RNN were built to extract the deep features of data, and one layer of the DNN was used to output the results. In both RNN layers, the tanh activation function and dropout were used to increase the fitting degree of the nonlinear model to the data and work for the model overfitting. The complete architecture of our RNN prediction research model is shown in Figure 5:

**Figure 5.**
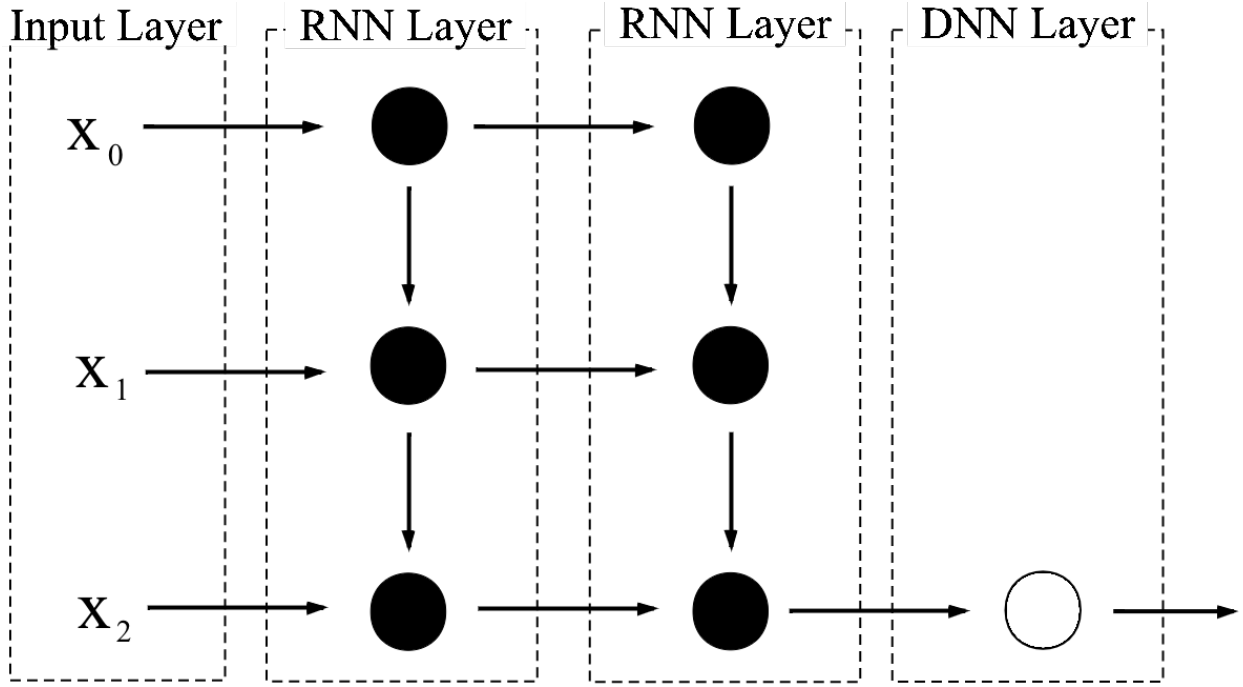
The architecture of the RNN. The number of hidden units weight in the first RNN layer was 100, the number of hidden units weight in the second RNN layer was 50 and the number of node in the DNN layer was 1.

## 3. Results

### 3.1. Prediction of Wuhan infection trend based on the SEIR model

According to the national health commission, by the end of March 6, the total number of confirmed cases was 80,653 [28]. Among them, 49,871 cases were confirmed in Wuhan, accounting for 60% of the total confirmed cases. The Chinese joint investigation report on novel coronavirus pneumonia (COVID - 19) [33] shows that the spread of the epidemic in China has distinct characteristics in the Wuhan and non-Wuhan areas of China. The newly infected cases in Wuhan were mainly original cases in Wuhan. As Wuhan is the traffic center of China and the epidemic spread rapidly during the Spring Festival travel rush, the newly infected cases in non-Wuhan areas were mainly imported from Wuhan. Since the newly added cases in Wuhan were mainly infected by original cases, the SEIR model was used to predict the epidemic trends in Wuhan.

To use the SEIR model, the contact rate *β*, incubation rate *σ*, recovery rate *γ* and other parameters needed to be estimated. The initial value of the susceptible population in Wuhan city was similar to that of the permanent resident population in Wuhan city. Because the incubation period of COVID-19 has been reported to be between 2 and 14 days, we chose the midpoint of 7 days. We used a recovery rate of 3% [34]. In the early stage of the outbreak, the number of infected people was small. The susceptible population S in the first 10 days in Wuhan was approximately the same as the population N on the same day in Wuhan. Therefore, S≈N. We obtain the new formula:

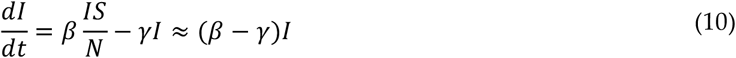

Finally, it is simplified to:

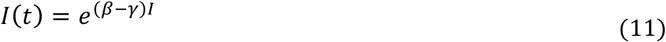

Based on the actual number of people infected in the first 10 days in Wuhan, this study estimated the initial *β* value to be 0.17.

Starting on February 12, the Hubei government changed the way it counted new confirmed cases. The number of new official diagnoses rose sharply as clinically diagnosed cases were included in new cases. Considering the change in the statistical method, this study corrected the initial parameters of the model appropriately to reduce the prediction error.

Based on these estimated parameters and the epidemiological data of Wuhan, the model parameters were fitted and optimized. The number of predicted results is shown in Figure 6, and the results are consistent with the actual situation. The SEIR model can predict the epidemic trend of the Wuhan area well.

**Figure 6.**
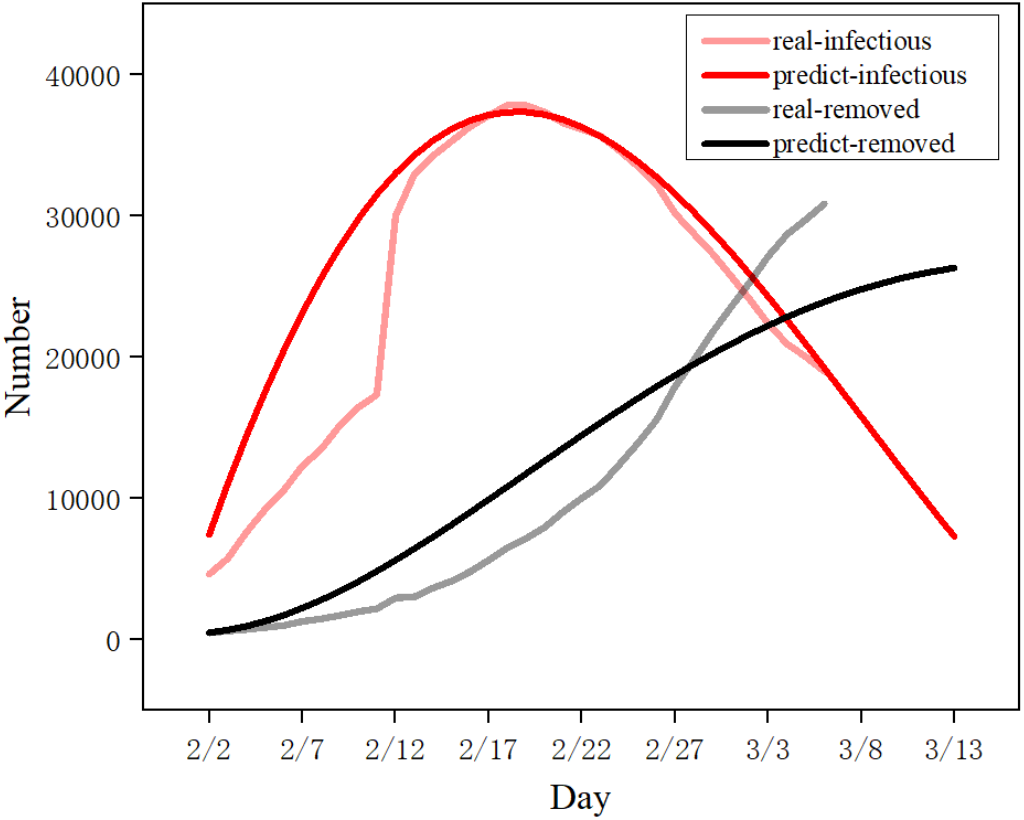
The SEIR model predicted the cumulative number of infections in Wuhan. Data from January 23 to March 3 were used to predict the cumulative number of infections in Wuhan over the next seven days.

### 3.2. Prediction of the non-Wuhan infection trend based on the DNN/RNN model

Since the newly infected cases in non-Wuhan areas were mainly imported from Wuhan, deep learning models were used to predict the epidemic trends in Wuhan. The number of new infections in non-Wuhan areas has a distinct character. The number of newly infected individuals in different provinces is shown in Figure 7:

**Figure 7.**
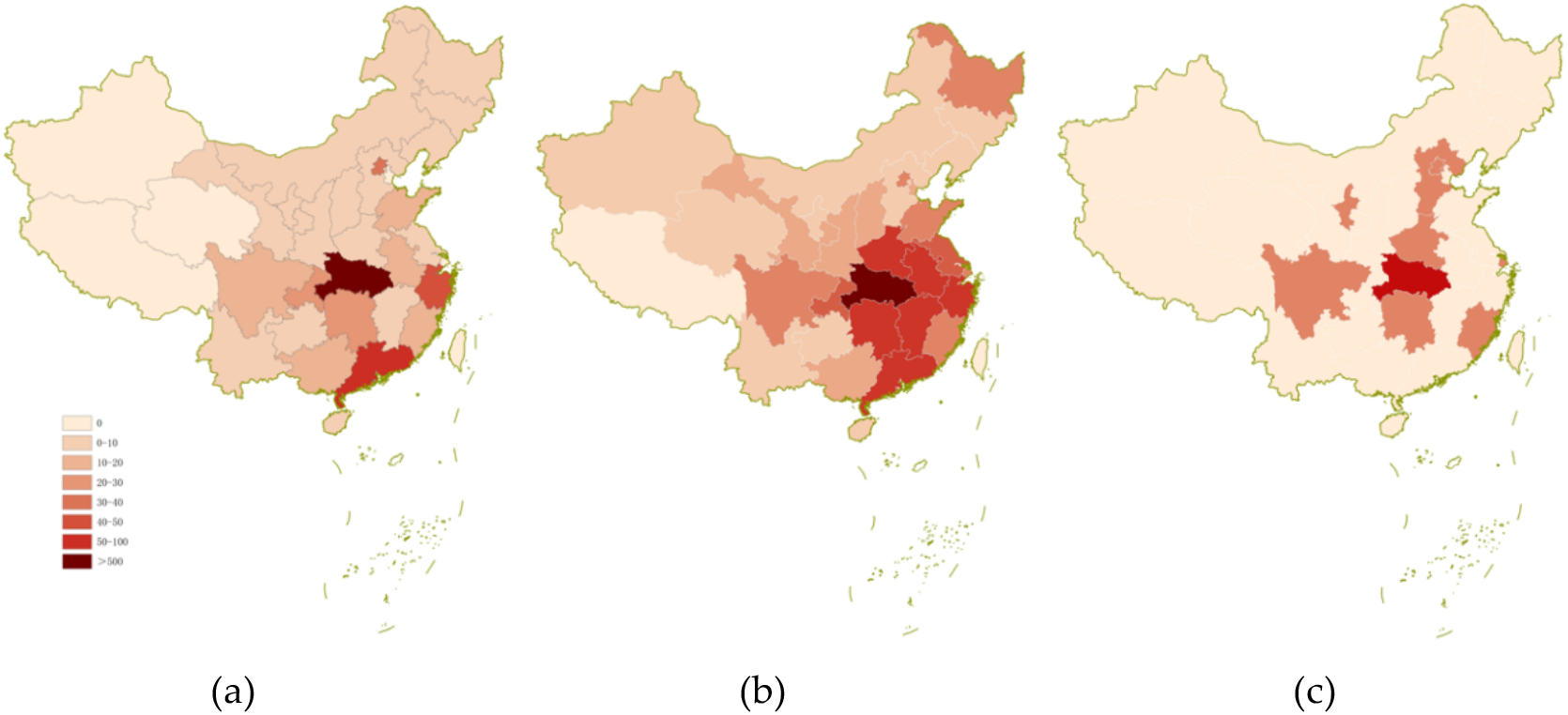
This figure shows the number of new infections in each province: (a) The number of new infections in each province on January 23; (b) The number of new infections in each province on February 2; (b) The number of new infections in each province on February 26.

The number of newly infected people in different provinces shows two main characteristics of the epidemic in non-Wuhan areas:

1. Wuhan was the radioactive center of the epidemic: the neighboring provinces of Hubei, including Hunan, Sichuan, Guangdong, Guangxi, Anhui, Zhejiang and Shandong, were seriously infected.
2. China’s supercities were the main focus of the epidemic: by February 2, in addition to the surrounding areas of Hubei, the newly infected cities in Beijing, Shanghai and Guangzhou were also serious. By February 26, as companies returned to work and provincial workers returned to central cities, new cases were reported in Beijing and Shanghai.

In view of the above characteristics, this study used the DNN/RNN model to predict the number of infected people in non-Wuhan areas of China.

The DNN model used nine types of data: epidemic data (the total number of confirmed cases in the previous day, the average number of newly confirmed cases in the past 3 days, and the number of existing cases in Wuhan), city data (local population density, per capita GDP, distance to Wuhan, average annual temperature, and average annual rainfall), and population migration data (the average of the migration population in Wuhan in the past 5 days). The RNN model used the values of these nine datasets over three consecutive days. Specific data are shown in Table 1:

**Table 1.**
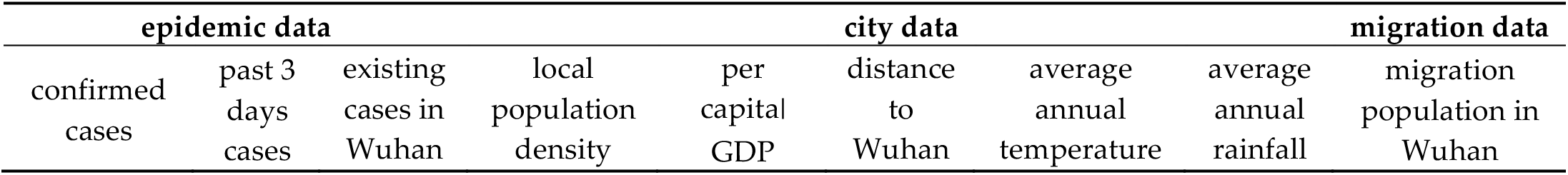
Data used by model DNN/RNN

Starting February 12, our model continuously predicted new infections in each province over the next three days. On March 6, we summarized the previous data and selected Beijing and Henan provinces with strong representation. The prediction effect of this DNN/RNN model is shown in Figure 8. It can be seen that the AI model with the population migration data can well predict the epidemic situation in non-Wuhan areas.

**Figure 8.**
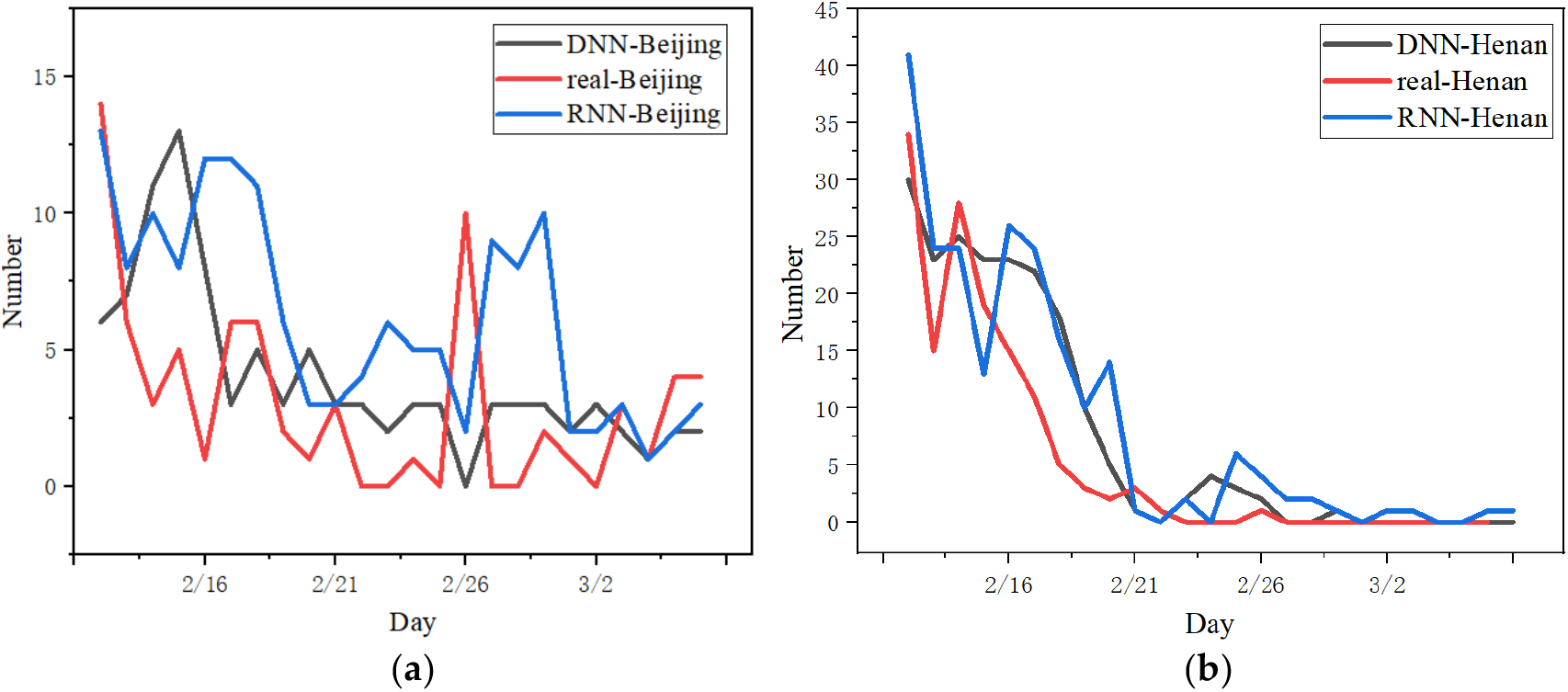
The number of newly confirmed cases predicted by the RNN/DNN model in each province: (a) results in Beijing; (b) results in Henan.

## 4. Discussion

Firstly, we used SEIR (Susceptible, Exposed, Infectious, and Removed) model fitting the epidemic trends of COVID-19 in Wuhan, China. And we used DNN(Deep Neural Network) and RNN(Recurrent Neural Network) model fitting the epidemic trends of COVID-19 in other parts of China.

Secondly, we used our model to predict the impact of closure of Wuhan on the the epidemic trends of COVID-19 in China.

Due to the outbreak, the Hubei provincial government launched a level-1 response to the public health emergency on January 23, 2020. The city’s buses, subways, ferries, coaches, airports and train stations were suspended. At the same time, all parts of the country also closed the city.

To explore the influence of Wuhan sealing on epidemic trends, this study attempted to simulate and predict the epidemic trends of open cities in all provinces. We used the data of Wuhan migration in 2019 to replace the data of Wuhan migration in 2020 to predict the number of people infected in non-Wuhan areas. We selected Beijing and Henan provinces with strong representativeness, and the prediction effect of the DNN model is shown in Figure 9.

**Figure 9.**
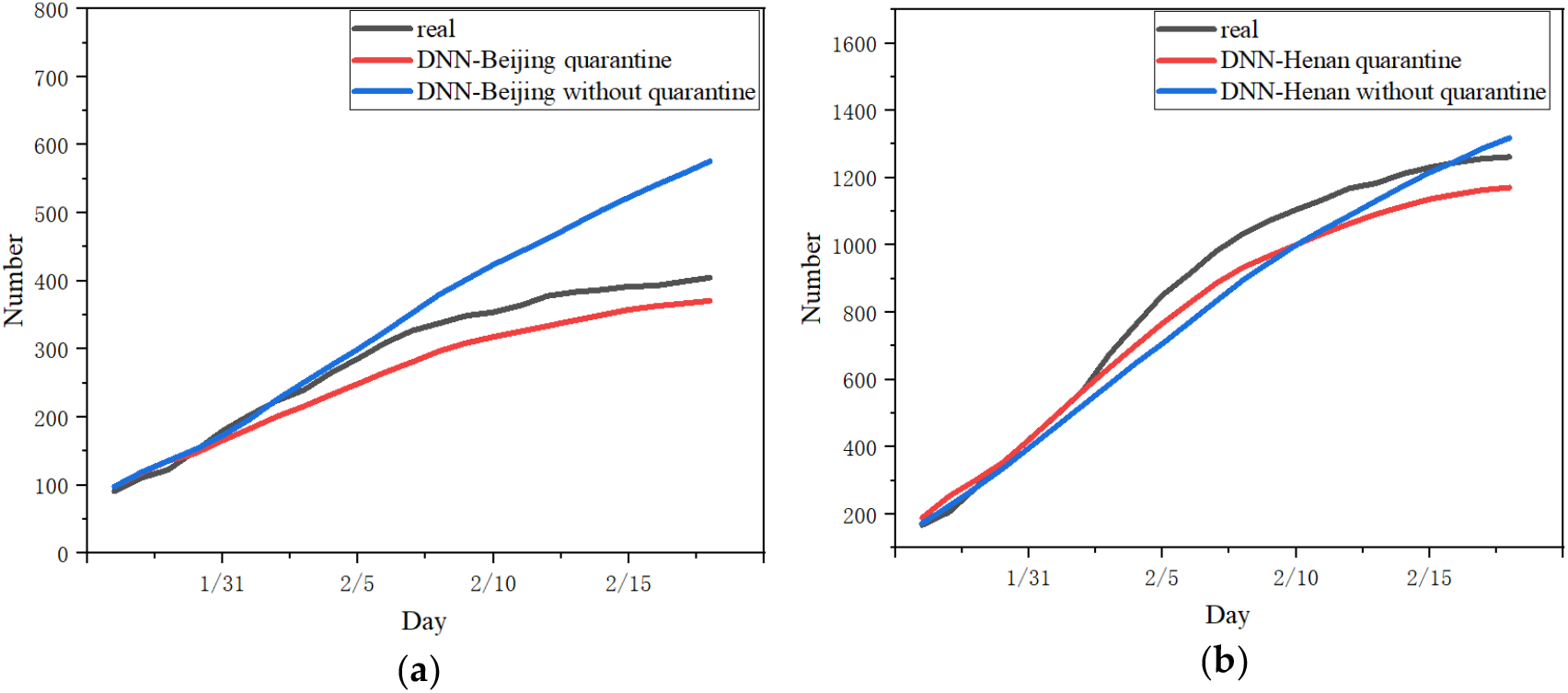
The DNN model was used to predict the total number of confirmed cases in provinces under the condition of city quarantine or not quarantine: (a) results in Beijing; (b) results in Henan.

Based on the model in this study, it is estimated that if Wuhan did not adopt city closure measures, for the vast majority of provinces such as Beijing, Chongqing and Guangdong, the cumulative number of increased infections in each province would have increased to 1.5-2.5 times within 3 weeks. This shows that the closure of the city had a great effect on inhibiting the further spread of the disease.

Additionally, we can see that the cumulative increase in the number of infections in different provinces did not occur in the early stages of the epidemic. Most provinces experienced a dramatic increase in the number of people infected within 3-7 days without closure. We believe this is due to the incubation period lasting for 2-14 days. If the city did not take measures to close the city, the new sources of infection in the city were mainly due to the latent period of the Wuhan migration population, and the outbreak was delayed by 3-7 days.

However, Henan Province and some other provinces with a smaller population did not conform to the above rule. In this study, it is believed that before Wuhan’s closure on January 23, Henan Province isolated the migrant population from Wuhan in a timely manner, so the impact of the Wuhan migrant population on the cumulative number of confirmed patients in Henan Province was small.

Our study has some limitations. Firstly, we built the model according to the conventional infection model, without considering the parameter fluctuation caused by the possible super disseminator and virus variation in the SEIR model.

Secondly, we use multiple data including epidemic data, urban data, migration data to predict the epidemic trends of COVID-19 in other parts of China, without considering the potential impact of other factors on COVID-19 in the DNN and RNN model.

Thirdly, in our models, our original parameters are based on previous studies and experience from SARS control. Besides, the data we using is based on the data before March 3, 2020. With the progression of COVID-19, the model parameters will change greatly because of more and more data.

## 5. Conclusions

In summary, this paper collected the COVID-19 data including the number of confirmed, cured and deaths from January 23 to March 6, 2020, combined with Baidu population migration data and relevant city data of the National Bureau of Statistics, to predict the number of infections in China. The SEIR model could well predict the epidemic situation in Wuhan, which was dominated by primary cases, and the AI model which added population migration data could well predict the epidemic situation in non-Wuhan areas in China with a large number of input infections. Additionally, this study estimated the influence of Wuhan closure on the epidemic trend. The results showed that the closure of Wuhan was an important measure to effectively inhibit the spread of COVID-19 in a large area, and it greatly reduced the number of infections in every part of China.

## Data Availability

1. The epidemic data that support the findings of this study are available in Ding Xiang Yuan (in Chinese) 2020, from http://www.dxy.cn/
2. The migration data that support the findings of this study are available in Baidu qianxi (in Chinese) 2020, from https://qianxi.baidu.com/
3. The city data that support the findings of this study are available in National Bureau of Statistics of China, Ovi interactive map and China Meteorological Administration, from http://www.dxy.cn/; http://www.gpsov.com/cn/main.php; http://www.cma.gov.cn/

https://qianxi.baidu.com/

http://data.stats.gov.cn

http://www.gpsov.com/cn/main.php

http://www.cma.gov.cn/

